# Watching the FIFA World Cup and Adult Sleep Quality: A Cross-Sectional Online Survey

**DOI:** 10.64898/2026.06.07.26355072

**Authors:** Fadi Aljamaan, Alaa A. Alanteet, Yazan Shaiah, Shereen Dasuqi, Mohammed A. Alarabi, Elshazaly Saeed, Serin M. Al-khatib, Abdallah A. Darweesh, Manan Raina, Khaled Saad, Khalid Alhasan, Ahmad S. BaHammam, Mohamad-Hani Temsah

**Author notes:** Correspondence: Mohamad-Hani Temsah, College of Medicine, King Saud University, Riyadh, Saudi Arabia. Both authors contributed equally to the manuscript (Fadi Aljamaan, Alaa A. Alanteet). **Conflicts of Interest** The authors declare no conflicts of interest.

## Abstract

Major international sporting events frequently impose exogenous demands that challenge adult circadian rhythms, often leading to the misalignment of sleep-wake cycles and social schedules. This cross-sectional study investigated the impact of the FIFA 2022 World Cup on adult sleep patterns to assess the prevalence and determinants of tournament-associated circadian disruption. Through an online survey, we captured data on sleep duration, timing, and subjective quality from a diverse adult population using Pittsburgh Sleep Quality Index (PSQI) score. The results indicate that 81.3% had high problematic sleep according to PSQI scores, while only 9% perceived that their sleep pattern was impacted by watching matches during the tournament. While 83.7% of the participants had low or mild anxiety according to GAD-7 scores, we found that GAD-7 scores correlated significantly with PSQI scores. Married participants had significantly lower PSQI scores (RR 0.856, *p =* .005), while those who reported that their sleep hours had changed during the tournament had significantly higher PSQI scores (1.180, *P-value* <0.001). Males reported a significantly high impact of the tournament on their sleep (OR 2.622, *P-value* <0.001). In conclusion, our data demonstrate a discrepancy between self-perception of sleep quality and self-rated assessment by PSQI scores, as well as the substantial impact of major international sporting events on adult sleep hygiene. The results provide data-driven insights helpful in evaluating potential circadian risks and informing public health strategies for major sporting events such as the FIFA world cup.

**Graphical Abstract:** 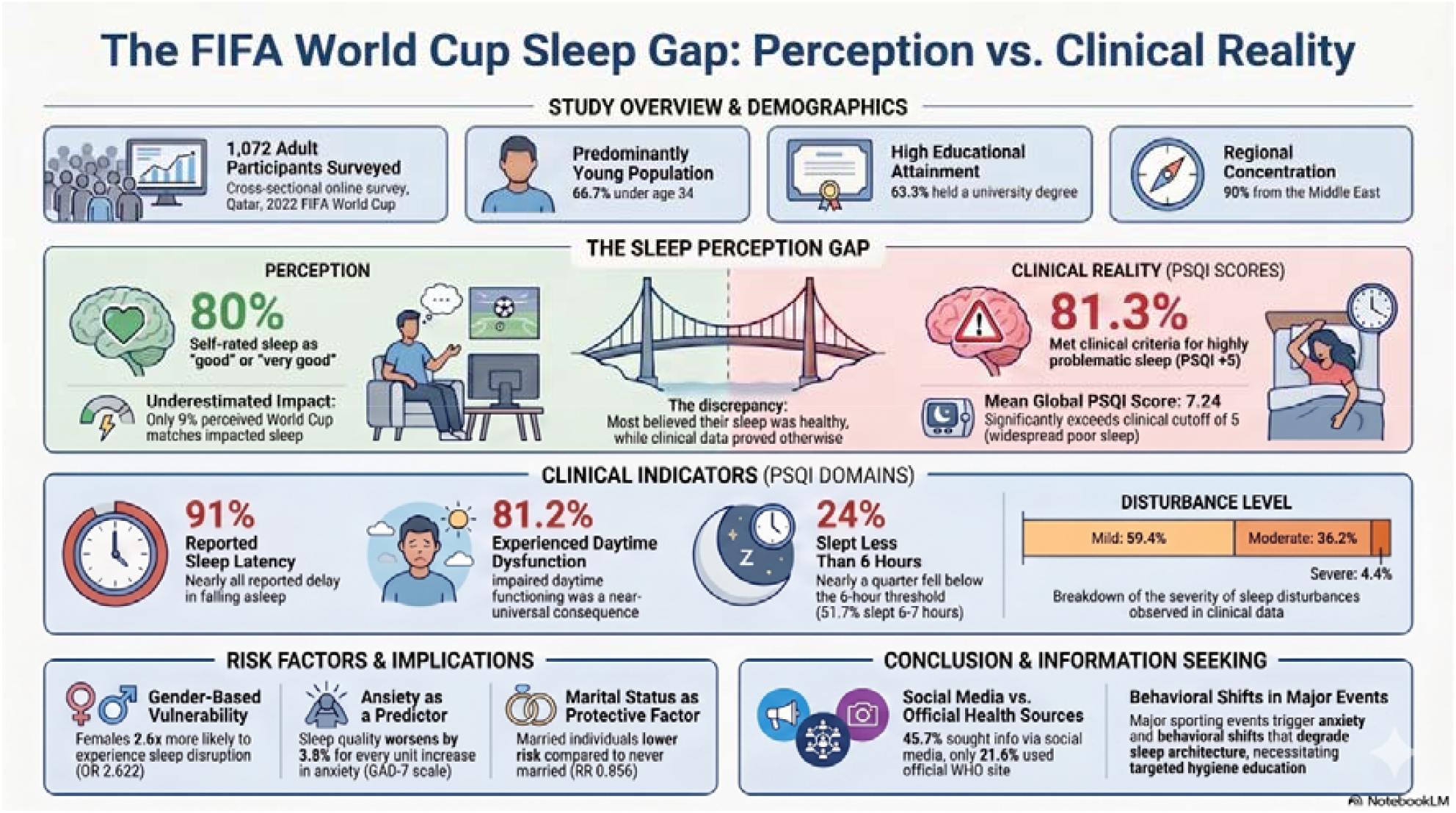

## Introduction

Sleep is a fundamental pillar of adult health, with its influence extending well beyond duration to encompass timing, regularity, continuity, and subjective quality. Emerging systematic evidence underscores that deviations from optimal sleep ranges, characterized by delayed and irregular schedules, are robustly associated with detrimental cardiometabolic, cognitive, and psychosocial sequelae [1]. A significant contributor to this misalignment is “social jetlag,” a phenomenon arising from the discrepancy between internal biological clocks and externally imposed social schedules, a condition that affects a large proportion of the adult population [2]. Major international social events, such as the FIFA World Cup, function as potent catalysts for this disruption; the congested schedules and late-evening fixtures inherent in elite sports often act as external stressors that force individuals to override biological sleep drives, exacerbating baseline social jetlag and extending adverse health consequences [3], [2].

Major international sporting events create unique exogenous pressures on circadian rhythm with mismatches between biological sleep-wake timing and social schedules.[4] The FIFA 2026 tournament, in particular, may introduce a multifaceted challenge among international football fans.[5] While the 2022 tournament compressed the sleep-deprivation window for many global viewers, the widespread distribution of North American venues necessitates a more granular, geofenced assessment of the interaction between match timing and local adult sleep health[6].

The FIFA 2022 World Cup functioned as a potent, naturalistic catalyst for circadian disruption, where irregular match-viewing habits frequently precipitated social jetlag and chronic sleep restriction[6]. These investigations clarify that the social exigencies of major sporting events act as reliable disruptors of established circadian rhythms, potentially exacerbating baseline social jetlag and extending adverse health consequences across diverse demographics [6].

The present study aimed to explore the sleep experiences of adults during the FIFA 2022 tournament to critically evaluate the prevalence and determinants of such disruptions. We hypothesized that watching FIFA matches would impact both sleep quality and quantity among adult fans. This observational, cross-sectional study hypothesized that watching FIFA 2022 matches would significantly impact adult sleep quality and quantity. By investigating these findings, we provide data-driven reflections intended to inform both public health strategies and individual sleep hygiene recommendations in anticipation of the upcoming FIFA 2026 World Cup in North America.[5]

## Methodology

### Study Design and Setting

This study was an observational, cross-sectional analysis of data collected during the FIFA World Cup 2022 in Qatar. Data collection was conducted between 27 November and 25 December 2022, coinciding with the tournament period. The study aimed to examine the perceived impact of watching FIFA World Cup matches on adult sleep quality, sleep quantity, and sleep timing, and to identify factors associated with poorer sleep during the tournament.

This analysis was based on the same survey framework used in our previous FIFA World Cup-related investigations, which examined tournament-associated sleep pattern changes and anxiety symptoms. For this analysis, we focused on adult participants and on variables relevant to sleep quality, sleep duration, sleep timing, social jetlag, match-viewing exposure, and anxiety symptoms.

### Participants and Recruitment

Participants were recruited using convenience and snowball sampling methods. Survey invitations were disseminated through widely used social media platforms, including WhatsApp, X, previously Twitter, and Facebook. Eligible participants were adults aged 18 years or older who were able to complete the questionnaire in Arabic or English and who followed the FIFA World Cup Qatar 2022 during the study period. And while the survey was primarily completed by participants from the Middle Easte, all interested adults were eligible to participate regardless of country of residence.

Before beginning the questionnaire, participants reviewed an electronic information sheet and provided digital informed consent. Participation was voluntary, and no identifying data were collected. Responses were anonymous, and confidentiality was maintained throughout the study period.

### Survey Instrument and Measures

The survey instrument was developed to capture sociodemographic characteristics, habitual adult sleep patterns, perceived sleep changes during the FIFA World Cup, match-viewing habits, and anxiety symptoms. An expert panel with clinical expertise in sleep medicine and psychiatry selected the Pittsburgh Sleep Quality Index (PSQI) [7] and the Generalized Anxiety Disorder-7 scale (GAD-7) [8] as the principal standardized instruments for assessing perceived sleep quality and anxiety symptoms during the tournament period.

Sociodemographic variables included age, gender, education, occupation, and residence. Sleep-related variables included habitual sleep duration, sleep timing on workdays and free days, perceived changes in sleep duration during the tournament, and PSQI sleep quality components. Social jetlag was operationalized consistently with our prior FIFA-related analyses as a delay in bedtime of one hour or more on free days compared with workdays [6]. FIFA-related sleep schedule change was assessed by comparing usual sleep timing with sleep timing during the tournament period.

Match-viewing exposure was assessed using self-reported frequency and duration of watching FIFA World Cup matches, as well as preferred method of following the matches. Anxiety symptoms were assessed using the GAD-7, a validated 7-item screening instrument measuring anxiety symptoms over the previous two weeks. Each item is scored from 0, “not at all”, to 3, “nearly every day”, yielding a total score ranging from 0 to 21, with higher scores indicating greater anxiety symptom severity.

### Statistical analysis

Continuous variables were described using means and standard deviations when they satisfied statistical normality assumptions. Continuous variables that violated normality assumptions were described using medians and interquartile ranges. The statistical normality assumption for metric variables was assessed using histograms and the Kolmogorov-Smirnov test. Categorical variables were described using frequencies and percentages. Multiple response dichotomies analysis was used to describe variables measured with more than one possible response option.

Internal consistency of the PSQI and GAD-7 was assessed using Cronbach’s alpha. The PSQI demonstrated acceptable reliability, with a Cronbach’s alpha of 0.744 across its seven main domains, indicating satisfactory internal consistency for measuring perceived sleep quality in this sample. The GAD-7 demonstrated excellent internal consistency, with a Cronbach’s alpha of 0.903 across its seven items, indicating a high degree of homogeneity among items measuring anxiety symptoms.

Bivariate Pearson’s correlation was used to assess correlations between metric variables. Collinearity between analyzed predictors was assessed using the Variance Inflation Factor (VIF) and Tolerance indices. Multivariable Generalized Linear Regression Analysis with a Gamma distribution was used to assess predictors of participants’ mean perceived sleep quality, as measured by the PSQI. This modeling approach was selected because the PSQI outcome showed a positively skewed distribution and because residuals from standard linear regression did not satisfy normality assumptions.

The overall model demonstrated adequate fit, with Akaike Information Criterion (AIC = 5078.1) and Bayesian Information Criterion (BIC = 5147.6), supporting the suitability of the Gamma specification for the analyzed data. Associations between independent predictor variables and the dependent sleep quality outcome were expressed as multivariable adjusted Risk Ratios, using exponentiated beta coefficients, with corresponding 95% confidence intervals.

All statistical analyses were performed using IBM SPSS Statistics version 21. The alpha significance level was set at 0.050.

### Ethical Considerations

The study protocol was reviewed and approved by the Institutional Review Board of King Saud University, Riyadh, Saudi Arabia (reference number: 19/0953/IRB), and was conducted in accordance with the Declaration of Helsinki. Participants provided voluntary electronic informed consent before completing the questionnaire and retained the right to withdraw at any stage without consequence. All data were securely stored and analyzed anonymously to maintain confidentiality throughout the research process.

## Results

A total of 1,072 participants were included in the study. The sample was relatively balanced by sex, with a slight predominance of females 52.6% compared to males 47.4%.

In terms of age distribution, the largest proportion of participants were young adults aged 18–24 years 31.3%, followed by those aged 25–34 years 25.4% and 35–44 years 24.5%. Older age groups were less represented, with 11.9% aged 45–54 years and only 6.9% aged 55 years or older, indicating a predominantly younger study population.

Regarding marital status, more than half of the participants were never married, 56.4%, while 43.6% reported being married. This aligns with the relatively young age structure of the sample.

Educational attainment was notably high, with most participants holding a university or postgraduate degree 83.3%, whereas only 16.7% had a high school education or less. This suggests a well-educated sample, which may have implications for health awareness and reporting behaviors.

With respect to employment status, 38.6% of participants were unemployed, representing the largest subgroup. This was followed by employed individuals (31.6%), and healthcare workers 25.4%, while a smaller proportion reported freelance work 4.4%. The relatively high proportion of unemployment may reflect contextual socioeconomic factors during the study period.

In terms of family structure, more than half of the participants reported having no children 56.4%, while 24.6% had 1–3 children and 17.2% had 4–6 children. Only a small minority (1.8%) reported having seven or more children. This distribution is consistent with the younger age profile observed in the sample.

Finally, most participants resided in Middle Eastern countries 90.0%, with only 10.0% from other regions, indicating that the findings are largely reflective of a Middle Eastern population context.

Based on the standardized scoring of the GAD-7 scale, more than half of the participants were classified as having low anxiety levels (55.2%, n = 592). Mild anxiety was observed in 28.5% (n = 306) of participants, while moderate and high anxiety levels were less accounting for 10.9% (n = 117) and 5.3% (n = 57), respectively, Figure.1.

**Figure 1:**
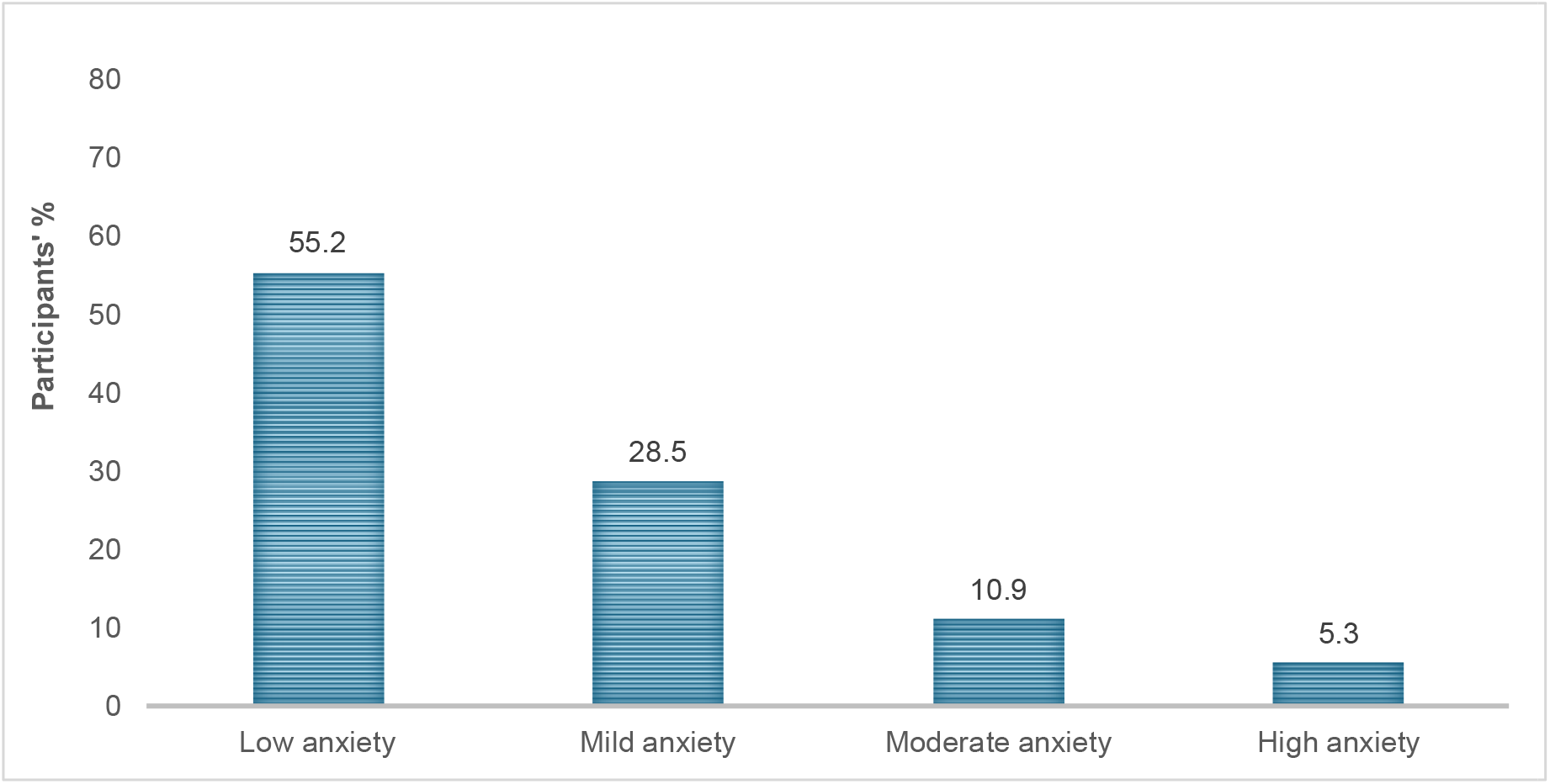
Participants’ Generalized Anxiety level as assessed by GAD-7 scale.

### Sleep Quality assessment by PSQI

The distribution of the Pittsburgh Sleep Quality Index (PSQI) components is presented in Table 3. Overall, participants reported varying levels of sleep quality across the seven PSQI domains.

**Table 1:** Descriptive characteristics of people’s sociodemographic characteristics, N=1072.

| **Variable** | **Frequency** | **Percentage** |
| --- | --- | --- |
| **Sex** |  |  |
| Female | 564 | 52.6 |
| Male | 508 | 47.4 |
| **Age group** |  |  |
| 18-24 years | 335 | 31.3 |
| 25-34 years | 272 | 25.4 |
| 35-44 years | 263 | 24.5 |
| 45-54 years | 128 | 11.9 |
| 55-64 years or older | 74 | 6.9 |
| **Marital state** |  |  |
| Never married | 605 | 56.4 |
| Ever married | 467 | 43.6 |
| **Educational Level** |  |  |
| High school or less | 179 | 16.7 |
| University or postgraduate degree | 893 | 83.3 |
| **Job status** |  |  |
| Freelance | 47 | 4.4 |
| Healthcare worker | 272 | 25.4 |
| Employed | 339 | 31.6 |
| Unemployed | 414 | 38.6 |
| **Number of children in the family** |  |  |
| None | 605 | 56.4 |
| 1-3 children | 264 | 24.6 |
| 4-6 children | 184 | 17.2 |
| >=7 children | 19 | 1.8 |
| **Region** |  |  |
| Middle eastern country | 965 | 90 |
| Other places | 107 | 10 |

**Table 3:** Descriptive analysis of people’s perceptions of sleep disturbance as per PSQI* main indicators.

| Variable | Frequency | Percentage |
| --- | --- | --- |
| **Component 1: Subjective sleep quality** |  |  |
| Very Good | 275 | 25.7 |
| Good | 604 | 56.3 |
| Bad | 170 | 15.9 |
| Very Bad | 23 | 2.1 |
| **Component 2: Sleep Latency** |  |  |
| None | 96 | 9 |
| Mild | 398 | 37.1 |
| Moderate | 397 | 37 |
| Severe | 181 | 16.9 |
| **Component 3: Sleep duration** |  |  |
| >7 hours | 257 | 24 |
| 6-7 hours | 554 | 51.7 |
| 5-6 hours | 171 | 16 |
| <5 hours | 90 | 8.4 |
| **Component 4: Sleep efficiency** |  |  |
| Very Good (>85%) | 648 | 60.4 |
| Good (75%-84%) | 187 | 17.4 |
| Poor (65-74%) | 124 | 11.6 |
| Ver poor (<=64%) | 113 | 10.5 |
| **Component 5: Sleep disturbance** |  |  |
| Mild | 637 | 59.4 |
| Moderate | 388 | 36.2 |
| Severe | 47 | 4.4 |
| **Component 6: Use of sleep medications** |  |  |
| None | 905 | 84.4 |
| Less than once/week | 92 | 8.6 |
| One or two per week | 50 | 4.7 |
| Three or more per week | 25 | 2.3 |
| **Component 7: Daytime dysfunction** |  |  |
| None | 201 | 18.8 |
| Mild | 500 | 46.6 |
| Moderate | 281 | 26.2 |
| Severe | 90 | 8.4 |
| **Global PSQI score, mean (SD)** |  | 7.24 (3.1) |
| **Global PSQI score level** |  |  |
| Low problematic sleep <5 | 201 | 18.8 |
| High problematic sleep | 871 | 81.3 |
| **Did your sleep hours change due to watching the World Cup matches?** |  |  |
| No or little | 975 | 91 |
| Yes (a lot) | 97 | 9 |

\* PSQI: Pittsburgh Sleep Quality Index

**Table 4:** Descriptive analysis of people’s used sources of sleep health information.

| Source | Frequency | Percentage |
| --- | --- | --- |
| Other sources | 608 | 59.4 |
| WHO website | 221 | 21.6 |
| Social media channels | 468 | 45.7 |
| Medical articles | 430 | 42 |

**Table 5:** Multivariable Generalized Linear with Gamma Regression Analysis of participants’ mean perceived Sleep Quality PSQI score.

| Variable | Multivariable adjusted Risk Ratio (RR) | RR 95% C.I. |  | p-value |
| --- | --- | --- | --- | --- |
|  |  | Lower | Upper |  |
| **(Intercept)** | 5.167 | 4.113 | 6.492 | <0.001 |
| **Sex** | 0.956 | .911 | 1.004 | .069 |
| **Male vs Female** |  |  |  |  |
| **Age group** | 1.002 | 0.976 | 1.030 | .869 |
| **Marital state** | 0.856 | 0.769 | 0.954 | .005 |
| **Ever married vs never married** |  |  |  |  |
| **Employment state** | 0.998 | 0.977 | 1.018 | .814 |
| **Educational Level** | 1.027 | 0.979 | 1.077 | .274 |
| **Location** | 1.033 | 0.951 | 1.121 | .444 |
| **International** |  |  |  |  |
| **Number Of kids in the family** | 1.048 | 0.981 | 1.120 | .165 |
| **Generalized Anxiety Disorder (GAD7) score** | 1.038 | 1.033 | 1.044 | <0.001 |
| **Source of information on sleep health** |  |  |  |  |
| **Other sources** | 0.951 | .904 | 1.000 | .049 |
| **WHO website** | 1.038 | 0.977 | 1.103 | .224 |
| **Social media channels** | 1.036 | 0.986 | 1.088 | .159 |
| **Did your sleep hours change due to watching the World Cup matches?** | 1.180 | 1.086 | 1.282 | <0.001 |
| **Yes vs No** |  |  |  |  |

*Dependent Variable: Mean perceived Global Pittsburgh Sleep Quality Index (PSQI) score. Estimation method: Maximum Likelihood with Gamma distribution Link. Model overall goodness-of fit criteria: AIC= 5078.1, BIC=5147.6.*

**Table 6:** Multivariable Binary Logistic Regression Analysis of participants with high FIFA tournament impact on their sleep hours.

| **Variable** | **Multivariable**<br>**adjusted Odds Ratio**<br>**(OR)** | **OR 95% C.I.** |  | **p-value** |
| --- | --- | --- | --- | --- |
|  |  | Lower | Upper |  |
| **Location** | 1.088 | .502 | 2.358 | .830 |
| **Gender** | 2.622 | 1.658 | 4.147 | <0.001 |
| **Male** |  |  |  |  |
| **Age group.** | 0.953 | 0.734 | 1.238 | .719 |
| **Educational Level** | 0.743 | 0.508 | 1.085 | .124 |
| **Employment state** | 1.170 | 0.958 | 1.429 | .124 |
| **Marital state** | 1.251 | 0.434 | 3.603 | .678 |
| **Ever**<br>**married vs**<br>**never**<br>**married** |  |  |  |  |
| **Number of**<br>**children in the**<br>**family** | 0.877 | 0.459 | 1.673 | .690 |
| **Generalized**<br>**Anxiety Disorder**<br>**(GAD7) score** | 0.993 | 0.947 | 1.042 | .789 |
| **Global Pittsburgh**<br>**Sleep Quality**<br>**Index PSQI score** | 1.163 | 1.078 | 1.255 | <0.001 |
| **Constant** | 0.042 |  |  | .005 |

*Dependent Variable: Did your sleep hours change due to watching the World Cup matches? No /Yes-A lot. Model overall significance $\chi^2(9)=39.9$ , p-value<0.001. Hosmer-Lemeshow G.O.F: $\chi^2(8)=10.84$ , p-value=0.211.*

Regarding subjective sleep quality, most participants rated their sleep as good 56.3% or very good 25.7% while a smaller proportion reported poor sleep quality bad: 15.9%; very bad 2.1%.

In terms of sleep latency, most participants reported some degree of delay in falling asleep, with 37.1% indicating mild latency and 37.0% reporting moderate latency. Severe sleep latency was reported by 16.9%, while only 9.0% experienced no latency difficulties, suggesting that delayed sleep onset was a common issue. For sleep duration, more than half of the participants reported sleeping 6–7 hours per night 51.7%, while 24.0% reported sleeping more than 7 hours. Short sleep duration was less common but still notable, with 16.0% sleeping 5–6 hours and 8.4% sleeping less than 5 hours.

With respect to sleep efficiency, most participants demonstrated good or very good sleep efficiency (60.4% and 17.4%, respectively), whereas 11.6% and 10.5% reported poor and very poor efficiency, respectively. This indicates that although many participants maintained adequate sleep efficiency, a substantial minority experienced reduced efficiency.

Regarding sleep disturbances, most participants reported mild disturbances 59.4%, while 36.2% experienced moderate disturbances and 4.4% reported severe disturbances, indicating that interruptions during sleep were relatively common.

The use of sleep medications was generally low, with most participants reporting no use 84.4%. Only small proportions reported occasional or frequent use, suggesting limited reliance on pharmacological sleep aids in this population. In terms of daytime dysfunction, nearly half of the participants reported mild dysfunction 46.6%, while 26.2% and 8.4% reported moderate and severe dysfunction, respectively. Only 18.8% reported no daytime dysfunction, indicating that impaired daytime functioning was relatively prevalent.

The mean global PSQI score was 7.24 (SD = 3.1), exceeding the recommended cutoff of ≥5, which indicates poor sleep quality. Consistent with this, most participants were classified as having high problematic sleep quality 81.3%, while only 18.8% were categorized as having low problematic sleep.

Most participants reported no or minimal changes of their sleep duration due to watching the tournament 91.0%, while a smaller proportion 9.0% reported substantial changes.

The analysis of participants’ reported sources of sleep health information revealed a diverse pattern of information-seeking behavior. As this was a multiple-response item, participants were able to select more than one source.

The most reported Sleep Health Information was other than WHO or social; media or medical articles sources 59.4% of participants, indicating that a substantial proportion relied on alternative or unspecified channels. Social media platforms were also widely used, reported by 45.7% of participants, followed closely by medical articles 42.0%, suggesting a moderate level of engagement with more formal or evidence-based information sources.

In contrast, the WHO website was less frequently utilized, with only 21.6% = 221) of participants reporting it as a source of information. This may reflect lower direct engagement with official global health platforms compared to more accessible or widely disseminated channels such as social media.

To assess the variables associated with sleep quality (PSQI score), a multivariable generalized linear regression model with a Gamma distribution and log link was conducted.

Among the sociodemographic variables, marital status was the only statistically significant predictor of sleep quality. Participants who were ever married had significantly lower PSQI scores compared to those who were never married (RR = 0.856, 95% CI: 0.769–0.954, p = .005), indicating relatively better sleep quality in this group. Other sociodemographic factors were not significantly associated with PSQI score.

The Generalized Anxiety Disorder (GAD-7) score was strongly and statistically significant predictor of sleep quality (RR = 1.038, 95% CI: 1.033–1.044, p < 0.001). This indicates that for each one-unit increase in anxiety score, the PSQI score increased by approximately 3.8%, reflecting poorer sleep quality.

Additionally, participants who reported that their sleep hours changed due to watching World Cup matches had significantly higher PSQI scores (RR = 1.180, 95% CI: 1.086–1.282, p < .001), corresponding to an approximate 18% increase in sleep disturbance (Figure 2).

**Figure 2:**
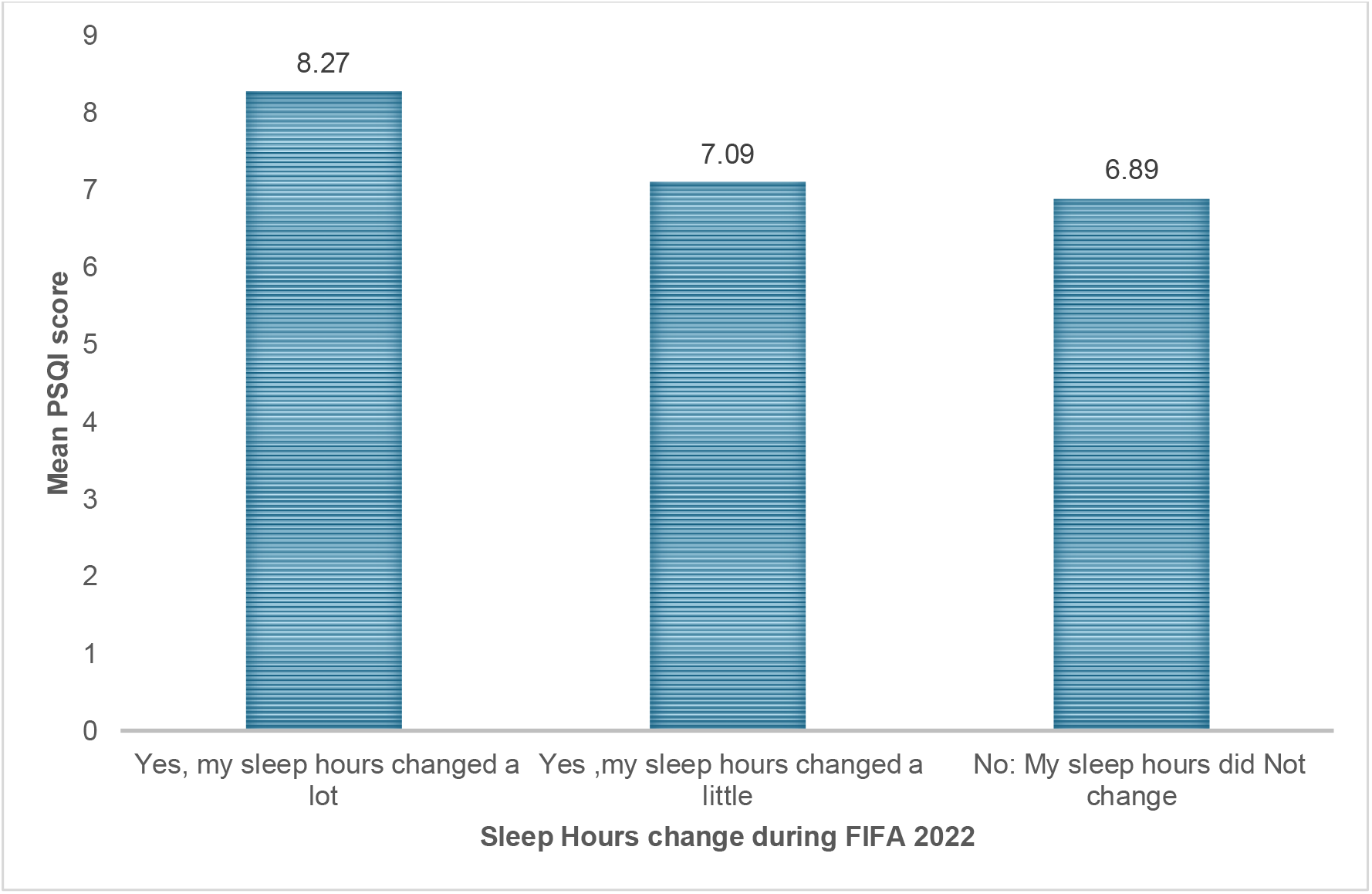
The association between participants’ sleep quality and the change in sleep hours during FIFA tournament. PSQI: Pittsburgh Sleep Quality Index.

Among the sources of sleep health information, reliance on other sources was marginally associated with lower PSQI scores (RR = 0.951, 95% CI: 0.904– 1.000, p = .049), suggesting a slight association with better sleep quality.

However, the use of other information sources were not significantly associated with sleep outcomes.

To assess the variables associated with participants’ substantial change in sleep hours during the FIFA World Cup 2022, a multivariable binary logistic regression analysis was conducted. The model was statistically significant, χ^2^(9) = 39.9, p < .001, indicating that the included predictors collectively contributed to distinguishing between participants who reported major changes in sleep hours and those who did not. The Hosmer–Lemeshow goodness-of-fit test was non-significant (χ^2^(8) = 10.84, p = .211), suggesting an adequate model fit between the data and the tested predictors set.

Male participants had significantly higher odds of reporting substantial changes in sleep hours compared to females (OR = 2.622, 95% CI: 1.658–4.147, p < .001).

The global PSQI score was also a significant predictor (OR = 1.163, 95% CI: 1.078–1.255, p < .001). Each one-unit increase in PSQI score was associated with a 16.3% increase in the odds of reporting major changes in sleep hours, suggesting that poorer baseline sleep quality predisposed individuals to greater susceptibility to behavioral sleep disruption during the event.

Other variables, including location, educational level, employment status, marital status and GAD-7 score were not significantly associated with changes in sleep hours after adjustment for other variables.

## Discussion

This study evaluates the intersection of psychological well-being and sleep health of majorly highly educated adult Middle Eastern cohort. Our objective is to assess their sleep quality during the FIFA World Cup season 2023. We uncovered a concerning discrepancy between subjective perceived sleep satisfaction and objective sleep quality assessed by PSQI score. A striking finding of this study is the high prevalence of poor global sleep quality, with 81.3% of the sample scoring above the PSQI clinical cutoff for highly problematic sleep[10]. Interestingly, this validated-scale measure strongly contrasted with participants’ subjective perceptions, as over 80% self-rated their sleep as “good” or “very good”. This discrepancy suggests a widespread lack of awareness regarding healthy sleep parameters; while individuals may feel they sleep well, a granular assessment reveals prevalent issues such as delayed sleep latency and daytime dysfunction.[11] This disconnect between self-assessment and clinical metrics mirrors patterns observed in other cohorts where reliance on subjective impressions frequently masks the underlying severity of sleep hygiene deficits [12].

Our data demonstrates a clear behavioral impact of the FIFA World Cup on sleep patterns, albeit concentrated within a specific subset of the population. Although only 9.0% of the overall sample reported substantial changes in sleep hours during the tournament, only 18% of the whole cohort were labelled to have low problematic sleep when assessed objectively by PSQI score.

Notably, male participants had more than twice the odds of experiencing marked sleep alterations during the tournament compared to females. While not directly measured in our source data, it is widely recognized in broader sociological and sports literature that male populations often exhibit higher engagement and viewership in major sporting events like the World Cup.[21] This cultural and behavioral difference may explain some of the sleep disruption differences. These observations necessitate a longitudinal comparative framework for the upcoming 2026 tournament, as prior investigations have consistently identified gender as a significant determinant in sleep disturbance and clinical insomnia prevalence [22,23]. Among the evaluated sociodemographic variables, marital status emerged as the primary significant predictor of baseline sleep quality. Specifically, an “ever married” status is associated with superior sleep outcomes underscoring the influence of social and familial stability [13]. This finding highlights the role of marital status in sleep hygiene [14]. Conversely, variables such as age, employment status, and education did not demonstrate a statistically significant influence on overall sleep quality in this sample. Although the high unemployment rate observed among these highly educated respondents likely reflects specific socioeconomic conditions, our data indicates that this factor did not independently drive sleep disruption, thereby underscoring the unique significance of marital status as a key sociodemographic determinant in this context.

Our regression analysis has shown that generalized anxiety was identified as a robust predictor of poor sleep quality [15]. For every one-unit increase in the GAD-7 anxiety score, PSQI score increased by approximately 3.8%. These results align with previous observations where psychological stress and inadequate sleep hygiene were found to be core drivers of mental health [16], further exacerbated by the potential for major sporting events to trigger psychological distress among fans. This vulnerability is often linked to the intensity of fan engagement and emotional investment, with research highlighting that such tournaments can disproportionately impact the mental well-being and sleep architecture of highly engaged populations, particularly among male demographics [17].

As global attention shifts toward the FIFA 2026 World Cup in North America, these insights serve as a foundational benchmark for future cross-continental comparisons, particularly as previous literature has demonstrated that tournament-associated anxiety and poor sleep quality are not localized phenomena but rather persistent global public health concerns [18]. These findings advocate for enhanced public health interventions that prioritize sleep awareness and anxiety management, particularly when major global events threaten to disrupt routine behaviors [19].

Furthermore, we observed that individuals who reported high impact of FIFA tournament on their sleep were significantly more susceptible to event-driven sleep disruption assessed by PSQI score. This relationship is underscored by our data, which indicates that every one-unit increase in the PSQI score correlates with a 16.3% increase in the odds of reporting major alterations to sleep duration during the tournament. Such findings suggest that individuals with problematic sleep according to their baseline PSQI score are vulnerable to behavioral shifts, including erratic scheduling—precipitated by late-night entertainment and major sporting events.

The study also highlights critical patterns in health information-seeking behaviors. Although this study was conducted in the period immediately following the COVID-19 pandemic—a time when public awareness of global health authorities was significantly elevated—participants still largely bypassed official global health platforms, with only 21.6% utilizing the World Health Organization website for sleep health information. Instead, reliance was heavily skewed towards social media and unspecified “other sources,” a trend corroborated by broader literature on post-pandemic health information acquisition [20]. Nonetheless, the high reliance on decentralized platforms underscores a communication gap; official health organizations may need to optimize their digital outreach and leverage social media more effectively to engage younger, educated demographics.

### Limitations and Strengths

This study offers a novel perspective as one of the first to explore sleep quality specifically in the context of the FIFA World Cup, employing validated assessment tools such as the Pittsburgh Sleep Quality Index [7] and the Generalized Anxiety Disorder-7 scale [21] to evaluate sleep quality and psychological distress. Nevertheless, these findings must be interpreted within the context of certain methodological limitations. The study cohort consisted predominantly of young, highly educated adults in Middle Eastern countries, which may constrain the generalizability of the results to older, less educated, or non-Middle Eastern populations. Additionally, the time zone of KSA where most participants resided was the same of the Qatar time zone where FIFA World Cup season 2022 was held, reliance on self-reported data introduces potential for recall or reporting bias, as subjective assessments of sleep metrics frequently differ from objective polysomnographic recordings [11]. Future investigations should prioritize the integration of wearable sensor technologies to capture longitudinal, objective sleep data across diverse international populations, thereby mitigating the biases inherent in cross-sectional, self-reported methodologies [22].

These limitations also provide opportunities for future research. Studies conducted during FIFA 2026 and other international sporting events should adopt longitudinal designs that assess sleep patterns before, during, and after the tournament to distinguish newly developed sleep disturbances from pre-existing sleep problems. Future work should also account for potential confounders, including prior sleep disorders, night-shift work, occupational schedules, recent life events, and post-pandemic lifestyle changes such as altered routines, reduced physical activity, and increased screen exposure. Including non-viewer control groups would strengthen causal inference by allowing comparisons between individuals exposed and not exposed to tournament viewing. In addition, future studies should assess participants’ level of football fandom, emotional engagement, match-viewing intensity, and time-zone differences, particularly during FIFA 2026, where matches will be held across multiple North American time zones. Integrating wearable sleep-monitoring technologies across diverse international populations may further enhance the objective assessment of sleep duration, timing, and quality during major sporting events.

## Conclusions

This study elucidates the complex interplay between psychological well-being, major sporting events, and sleep architecture. Our findings reveal a marked disconnect between perceived and actual sleep quality, which is significantly exacerbated by anxiety and behavioral disruptions triggered by global events. Consequently, public health initiatives must prioritize sleep hygiene education and targeted anxiety management, particularly during high-profile tournaments that threaten to derail established routines, with a specific focus on vulnerable demographics. Furthermore, to better understand the causality between fan engagement intensity and sleep health, longitudinal research is essential. Future studies conducted during upcoming global spectacles, such as the FIFA 2026 tournament and similar large-scale sporting events, are urgently needed to delineate the long-term impacts of these events on population-level sleep patterns and mental well-being.

## Data Availability

All data produced in the present study are available upon reasonable request to the corresponding author

## Acknowledgements

The authors acknowledge the use of generative artificial intelligence (AI), specifically ChatGPT (OpenAI) was utilized to refine and polish the manuscript’s language and phrasing, and NotebookLM (Google) and NanoBanana 2 were employed to generate the initial concepts and layout for the Graphical Abstract. Following the AI-assisted generation, all authors thoroughly reviewed, verified, and edited the final outputs, and take full responsibility for the content, integrity, and accuracy of the final version of the manuscript and its accompanying figures.

